# Suicide and depression in former contact sports participants: population-based cohort study, systematic review, and meta-analysis

**DOI:** 10.1101/2022.11.11.22282212

**Authors:** G. David Batty, Philipp Frank, Urho M. Kujala, Seppo J. Sarna, Jaakko Kaprio

**Affiliations:** Department of Epidemiology and Public Health, University College London, London, UK; Faculty of Sport and Health Sciences, University of Jyväskylä, Jyväskylä, Finland; Department of Public Health, University of Helsinki, Helsinki, Finland; Institute for Molecular Medicine FIMM, University of Helsinki, Helsinki, Finland

**Author notes:** Corresponding author: David Batty, Department of Epidemiology and Public Health, University College London, 1-19 Torrington Place, London, UK, WC1E 6BT. E. Contributions: GDB generated the idea for the paper, formulated the plan for analyses of the cohort data, conducted the literature search for the systematic review, extracted results, prepared tables and figures, and drafted the manuscript. PF conducted the literature search for the systematic review, carried out the meta-analysis, prepared figures, and edited the manuscript. UMK and SJS initiated the cohort study, designed data collection, and edited the manuscript. JK designed data collection in the cohort study, formulated the plan for analyses of cohort data, conducted those analyses, and edited the manuscript. Data sharing: Interested parties should contact UMK and SJS regarding access to the Finnish cohort study data.

## Abstract

**Background:** Traumatic brain injury is associated with the future risk of depression and suicide, and this raises the possibility that former participants in sports characterised by low intensity repetitive head impact may also subsequently experience an increased burden of these mental health outcomes. Using new data from a cohort study integrated into a meta-analysis of the current evidence, we compared the occurrence of depression and suicide in former contact sports athletes against general population controls.

**Methods:** The cohort study comprised 2004 retired male athletes who had competed internationally on an amateur basis for Finland between 1920 and 1965, and 1385 age-equivalent male general population controls. Former contact sports participants were drawn from soccer, boxing, or Olympic-style (non-professional) wrestling. During follow-up, cases of major depressive disorder and suicide were captured via linkage to mortality and hospitalisation registries. In a PROSPERO-registered (CRD42022352780) systematic review, we searched PubMed and Embase from their inception to October 31 2022 for reports of cohort studies of contact sports participation and later risk of depression and suicide that were published in English and reported standard estimates of association and variance. Study-specific estimates were aggregated using random-effect meta-analysis. The Newcastle-Ottawa Scale was used to appraise the quality of each study.

**Findings:** In analyses of the Finnish cohort data, up to 45 years of health surveillance gave rise to 131 hospitalisations for major depressive disorder and 61 suicides. In survival analyses (hazard ratio [95% confidence interval]) adjusted for age and socioeconomic status, former boxers (depression: 1.43 [0.73, 2.78]; suicide: 1.75 [0.64, 4.38]), wrestlers (depression: 0.94 [0.44, 2.00]; suicide: 1.60 [0.64, 3.99]), and soccer players (depression: 0.62 [0.26, 1.48]); suicide: 0.50 [0.11, 2.16]) did not have statistically significantly different rates of major depressive disorder or suicide relative to general population controls. The systematic review identified 463 potentially eligible published articles, of which 7 met inclusion criteria. All studies sampled men and 3 were evaluated as being of high quality. After aggregating results from these retrieved studies with new data from the cohort study, we found that retired soccer players appeared to have a somewhat lower risk of depression (summary risk ratio: 0.71 [95% 0.54, 0.93]) relative to general population controls, while the rate of suicide was essentially the same (0.70 [0.40, 1.23]). Past participation in American football was associated with some protection against suicide (0.58 [0.43, 0.80]) but there were insufficient studies of depression for aggregation. All studies showed directionally consistent relationships and there was no indication of inter-study heterogeneity (I^2^=0%).

**Interpretation:** Based on a small cluster of studies exclusively comprising men, retired soccer players had a lower rate of later depression, and former American football players had a lower risk of suicide. Whether these findings are generalisable to women requires testing.

**Funding:** None.

**Research in context:** *Evidence before this study:* A series of cohort studies suggest that brain injury serious enough to necessitate hospitalisation is associated with elevated rates of later depression and suicide. This raises the possibility that former participants in sports characterised by low intensity repetitive head impact, who have an increased risk of other mental health disorders in later life such as dementia, may also experience an increased burden of depression and suicide. Searching PubMed and Embase using terms for specific contact sports (e.g., ‘boxing’, ‘martial’, ‘wrestling’, ‘football’, ‘soccer’, ‘hockey’, ‘rugby’), depression and suicide (e.g., depression, dysthymic; suicide) revealed relevant studies in former athletes from American football, soccer, and rugby union but no evidence for boxing, wrestling, or other contact sports. Overall, there was a suggestion of mixed results and an absence of a quantitative synthesis of findings for depression.

*Added value of this study:* In the first cohort study to simultaneously examine the risk of depression and suicide across multiple contact sports, there was no convincing evidence that retired boxers, wrestlers, or soccer player had a different rate of these health outcomes than the general population. After incorporating these new results into a meta-analysis, former soccer players had a lower risk of depression but there was no clear link with suicide. Retired American footballers appeared to experience lower suicide rates at follow-up.

*Implications of all the available evidence:* Counter to the apparent impact of traumatic brain injury, a background in contact sports was not associated with elevated rates of depression or suicide. Indeed, former soccer athletes (depression) and American football players (suicide) seemed to experience some protection against these health outcomes. The existing evidence base is, however, hampered by an absence of studies of women, and is modest in scale and narrow in scope, currently not including several popular contact sports.

## Introduction

A series of large-scale cohort studies demonstrate a markedly higher rate of suicide and depression in individuals with a history of traumatic brain injury severe enough to require hospitalisation relative to unaffected population controls.^1,2^ In the more comprehensive investigations, these effects were independent of confounding factors, including comorbidities.^3^ These observations raise the possibility that a history of involvement in sports characterised by repetitive low-level head impact, such as boxing, soccer, and American football, might be linked to the development of depression and suicide, as seems to be the case for other mental health disorders such as dementia and Alzheimer’s disease.^4^

Much of the evidence for contact sports having an impact on depression and suicide stems from case reports of select athlete samples where a post-mortem diagnosis of chronic traumatic encephalopathy, formerly termed dementia pugilistica, is reportedly accompanied by depression, suicide, or aggressive behaviours.^5,6^ While these data may be hypothesis-generating, any potential link between past participation in contact sports, depression, and suicide can be examined more robustly in cohort studies in which the long-term health experience of retired athletes is compared with unaffected population controls.^7^ Such studies are rare however, and seem to reveal discordant findings such that a lower incidence of depression in retired soccer players^8^ and suicide in former American football professionals^9^ has been reported relative to unexposed individuals, while in other studies, no such group differences were evident.^10,11^ It is plausible that the contrasting profile of head impacts across different contact sports may account for different patterns of disease risk but such cross-sport comparisons are currently lacking.^12^

We address these uncertainties in two ways. We first report new results from a cohort of retired amateur athletes representing an array of elite-level sporting backgrounds, and then integrate these findings into a meta-analysis based on a systematic review of the available literature. To the best of our knowledge, there is one existing meta-analysis of suicide in former contact sports^13^ but we were unable to identify the same for depression. That global participation in soccer – estimated at more than a quarter of a billion by its governing body^14^ – is seemingly the highest of any sport, and programmes of American football are long-established in educational settings,^15^ means that a link between a background in these activities and depression or suicide may have public health significance.

## Methods

### Cohort of Finnish former elite athletes

This cohort study was initiated in 1978 to examine the relationship between participation in different elite-level sports and long-term health.^16-18^ The sample was generated from 2613 Finnish male former elite amateur athletes (640 Olympians) who competed internationally between 1920 and 1965. Identified using a combination of sports yearbooks, association records, and periodicals, these individuals represent an array of sporting backgrounds. Data collection was approved by the ethics committee of the Hospital Districts of Helsinki and Uusimaa, and all participants consented. The reporting of this cohort study conforms to the Strengthening the Reporting of Observational Studies in Epidemiology (STROBE) Statement of guidelines for reporting observational studies.^19^

#### Derivation of exposed and unexposed groups

Based on the occurrence of head injury,^20-22^ sports were denoted as contact (soccer, boxing, wrestling, ice hockey, and basketball) or non-contact (track and field, cross-country skiing, and weight-lifting). Individual contact sports were then disaggregated as the numbers of cases of depression and suicide allowed. Thus, separate analyses were possible for former soccer players, boxers, and wrestlers (non-professional or freestyle/Greco-Roman), while retired athletes from the sports of ice hockey and basketball players were combined into a ‘other’ contact sports category. A general population-based (unexposed) comparison group comprising age-equivalent healthy men was generated using records from a population-wide medical examination routinely administered to all Finnish males at around 20 years of age as part of national service (N=1712). Participants in non-contact sports represented a further comparison group unexposed to head impact. Using data from a 1985-administered questionnaire in combination with those extracted from the Finnish Central Population Registry, we generated a variable for longest held job, our indicator of socioeconomic status.^23^

#### Ascertainment of depression, depression ‘caseness’, and suicide

Health surveillance of study members began upon initiation of nationwide health registries in 1970 when the average age of the athlete group was 45.4 years (controls 44.3 years). Study members were linked to death (suicide) and hospitalisation (depression and suicide) records. Major depressive disorder was coded according to the International Classification of Disease (ICD) version eight (29600, 29620, 30040, 30041), nine (2961, 2968A, 3004A), or ten (F32-34). The ICD codes used to denote suicide were E950-E959 (version eight and nine), or X60-X84 (version ten).

As part of a mailed questionnaire resurvey in 1995, surviving study members completed the Brief Symptom Inventory,^24^ a 53 item scale of psychological distress. For each of the 6 items that comprise the depression subscale, respondents used a 5 point continuum (0-4) to indicate the extent to which they had been concerned by suicidal ideation, loneliness, or a lack of interest in usual activities in the prior week (total 0-24). ^24^ We used a score of ≥11 to denote depression ‘caseness’. This threshold was strongly associated with subsequent risk of hospitalisation for major depressive disorder (age-adjusted odds ratio: 6.41 [95% CI 3.82, 10.75]) and suicide (age-adjusted hazard ratio: 7.58 [95% CI 2.33, 24.69] in the expected direction, suggesting some predictive validity.

### Systematic review and meta-analysis

#### Search strategy and study selection

This PROSPERO-registered (CRD42022352780) systematic review and meta-analysis is reported in accordance with the guidelines for Preferred Reporting Items for Systematic reviews and Meta-Analyses (PRISMA).^25^ We identified relevant literature by searching PubMed (Medline) and Embase databases between their inception and October 31 2022. We used combinations of free text and controlled terms in 2 categories (supplemental box 1): the exposure (e.g., specific sports such as boxing, soccer, martial arts, and rugby), and the outcome (e.g., depression and suicide). We also scrutinised the reference sections of retrieved articles for additional publications.

We included a published paper if it fulfilled the following criteria: utilised a cohort study design; a comparison of depression and/or suicide occurrence was made between a group of former contact sports athletes and unexposed (general population) or lesser-exposed (former athletes from non-contact sports) controls; the identification of former participants in contact sports was records-based (e.g., pension or union registers, association or school/college yearbooks) rather than being self-declared; standard estimates of association (e.g., relative risk, odds ratios, hazard ratios) and variance (e.g., confidence interval, standard error) were reported or could be calculated based on the data provided; published in a peer-reviewed journal; and published in English. Two authors (GDB and PF) independently screened the identified records first by title, then abstract, and, if necessary, the full paper. Any discrepancies were resolved on discussion.

#### Extraction of results and assessment of study quality

Where available, a range of characteristics were extracted from each publication, including the name of the lead author, publication year, country of sample population, number exposed and unexposed participants, number of events, and effects estimates for both minimally- and multivariable-adjusted results. Study authors were contacted when clarification was required.

We used the Newcastle-Ottawa Scale to appraise the quality of each study (supplemental table 1).^26^ Comprising eight domains, including the comprehensiveness of exposure and outcome ascertainment and adequacy of the period of health surveillance, a maximum of 1 point (range: 0-1) is allocated for each domain (total 9) with a higher score denoting better quality. Studies with a score of ≥7 were regarded as high grade.

### Statistical analyses

In individual-participant analyses of the Finnish cohort study, after exclusion of study members owing to record-linkage failure and death prior to the beginning of follow-up, the main analytical sample comprised 3389 men (2004 former athletes, 1385 population controls). Event surveillance was from 1^st^ January 1970 until the occurrence of a depression or suicide event or the end of the surveillance period (December 31, 2015) – whichever came first. Having ascertained that the proportional hazards assumption had not been violated, we used Cox regression to compute hazard ratios with accompanying 95% confidence intervals to summarise the relationship of a background in contact sports with later risk of depression and suicide.^27^ Age was used as the time covariate. In the main analysis of depression scores from the Brief Symptom Inventory, we used linear regression to compute beta coefficients with accompanying 95% confidence intervals. For inclusion of these symptoms scores in the meta-analysis where papers reported odds or hazard ratios, we categorised the depressions scores according to caseness, and used logistic regression to compute odds ratios and 95% confidence intervals.

For the meta-analysis, we pooled the results from analyses of the Finnish cohort alongside published study-specific estimates using a random effects meta-analysis,^28^ an approach which incorporates the heterogeneity of effects in the computation of their aggregation. An I^2^ statistic was computed to summarise the heterogeneity in estimates across studies. Individual-participant analyses were performed using Stata 15 (StataCorp, College Station, TX), and the Meta-aanlysis was conducted using R.

### Role of the funding source

This report had no direct funding. All authors had final responsibility for the decision to submit for publication.

## Results

### Finnish cohort study

In analyses of the Finnish cohort data, up to 45 years of health event surveillance in an analytical sample of 3389 men gave rise to 131 hospitalisations for major depressive disorder, and 61 suicides (20 attempts, 41 deaths). A subgroup of 1419 men responded to the Brief Symptom Inventory. Taken together, there was little clear evidence of an association between a history of contact sports participation and later risk of major depressive disorder (table 1). Thus, while retired boxers seemed to experience some non-significantly raised risk of depression (age- and socioeconomic status-adjusted hazard ratios [95% confidence interval]: 1.43 [0.73, 2.78]), past participation in wrestling was unrelated to this mental health outcome (0.94 [0.44, 2.00]). Erstwhile soccer players (0.62 [0.26, 1.48]) and other collision sports participants (0.75 [0.34, 1.63]) had non-significantly lower rates of major depressive disorder relative to general population controls. Based on the subgroup of participants who responded to questionnaire enquiries about depression symptoms, there was a lower depression score amongst one-time wrestlers (age- and socioeconomic status -adjusted beta coefficient [95% confidence interval]: -1.04 [-1.92, -0.17]). A lower depression symptom score was also apparent, however, in athletes who formerly engaged in non-collision sports (-0.69 [-1.22, -0.15]). Analyses in which we computed odds ratios based on depression caseness for the purposes of inclusion in the meta-analysis produced a similar pattern of results (supplemental table 2).

**Table 1.**
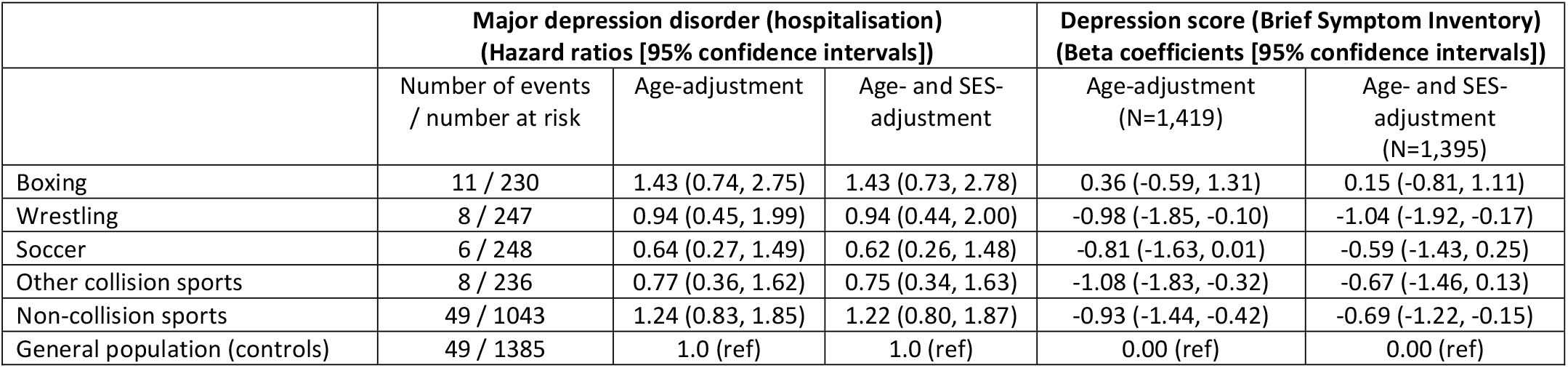
Association of participation in contact sports with hospitalisation for major depressive disorder and self-reported depression score: Finnish cohort study.

In the analyses of suicide events in the Finnish cohort study (table 2), there was some suggestion of an elevated risk in retired boxers (age- and socioeconomic status -adjusted hazard ratio: 1.75 [95% CI 0.64, 4.38]), as there was for former wrestlers (1.60 [0.64, 3.99]); again, however, these effect estimates did not attain statistical significance. Confidence intervals were also wide on occasion, indicating low statistical power owing to a small number of events.

**Table 2.** Hazard ratios (95% confidence intervals) for the association of participation in contact sports with suicide: Finnish cohort study.

|  | Suicide |  |  |
| --- | --- | --- | --- |
|  | Number of events / number at risk | Age-adjustment | Age- and SES-adjustment |
| Boxing | 6/230 | 1.57 (0.65, 3.84) | 1.75 (0.64, 4.38) |
| Wrestling | 6/247 | 1.44 (0.59, 3.53) | 1.60 (0.64, 3.99) |
| Soccer | 2/248 | 0.43 (0.10, 1.83) | 0.50 (0.11, 2.16) |
| Other collision sports | 3/236 | 0.54 (0.16, 1.79) | 0.61 (0.17, 2.13) |
| Non-collision sports | 19/1044 | 1.00 (0.55, 1.82) | 1.14 (0.60, 2.20) |
| General population (controls) | 25/1386 | 1.0 (ref) | 1.0 (ref) |

Lastly, we carried out some sensitivity analyses. The somewhat lower rates of depression and suicide in former contact sports athletes could be ascribed to their more favourable risk factor profile. Thus, relative to the general population, post-retirement, athletes tended to have a lower prevalence of smoking, heavy alcohol intake, and socioeconomic deprivation, and it is these factors rather than their status as former contact sports participants that lowers their risk of suicide and depression. The questionnaire mailed in 1985 captured these covariates and in analyses in which we collapsed all contact sports participants into a single group to preserve statistical power (68 cases of major depressive disorder in follow-up of 1897 men, and 20 cases of suicide in 1913 men from 1985), our conclusions were unchanged.

### Systematic review and meta-analysis

Our systematic review retrieved 463 potentially eligible published articles of which 7 met the inclusion criteria (figure 1).^8–11,15,29,30^ The characteristics of the included cohort studies are summarised in table 3. Three featured depression only as the outcome of interest,^11,15,30^ 3 reported on only suicide,^9,10,29^ and 1 captured both endpoints.^8^ All studies exclusively comprised men and, bar two,^15,30^ sampled former professional athletes. The number of events ranged from 21^30^ to 38^8^ for depression, and 8^10^ to 19^8^ for suicide. Of the 7 retrieved studies, 3 were evaluated as being of high quality.^8,9,29^

**Table 3.**
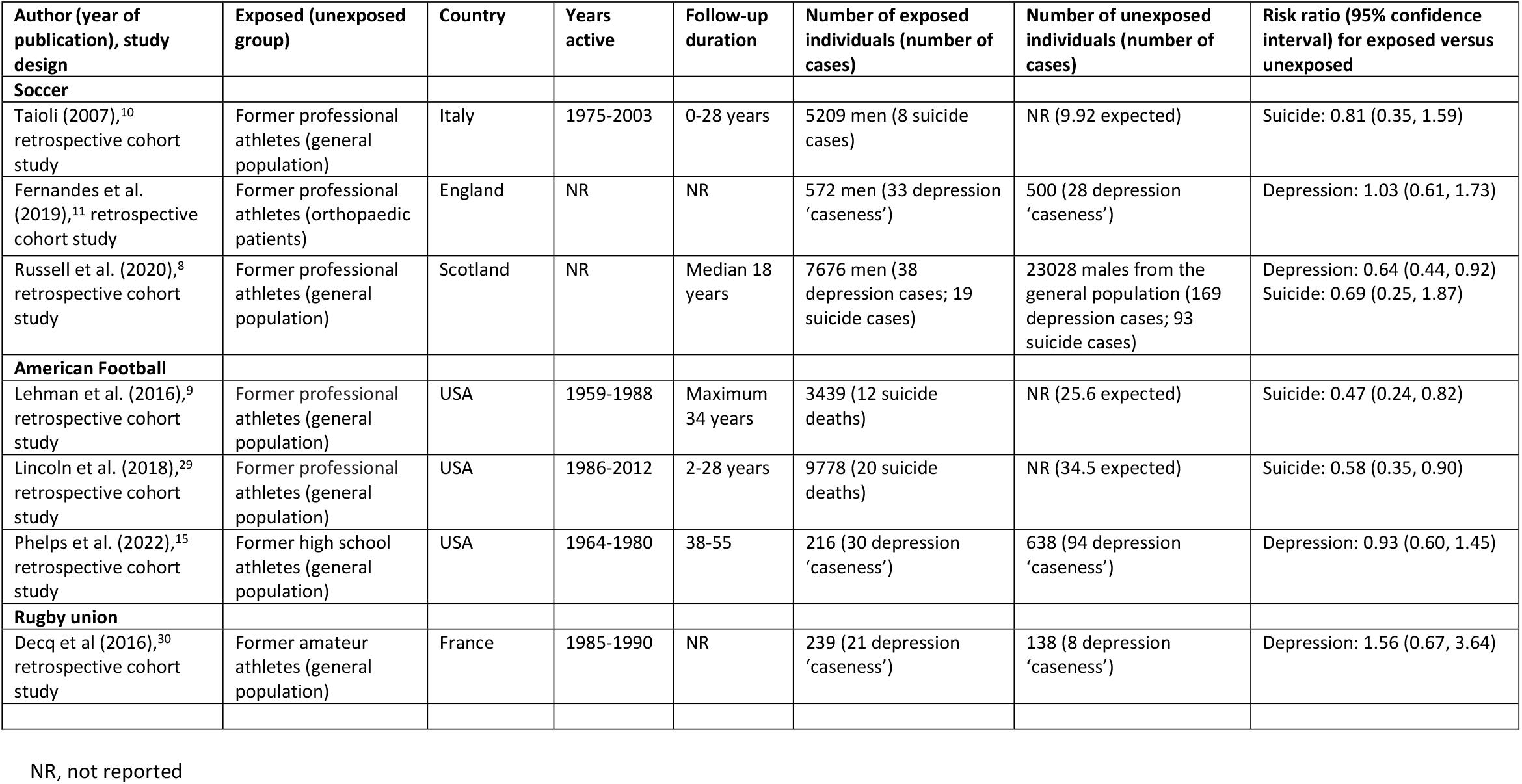
Former participation in contact sports and risk of depression and suicide: Characteristics of studies included in the meta-analysis.

| Author (year of publication), study design | Exposed (unexposed group) | Country | Years active | Follow-up duration | Number of exposed individuals (number of cases) | Number of unexposed individuals (number of cases) | Risk ratio (95% confidence interval) for exposed versus unexposed |
| --- | --- | --- | --- | --- | --- | --- | --- |
| **Soccer** |  |  |  |  |  |  |  |
| Taioli (2007),<sup>10</sup> retrospective cohort study | Former professional athletes (general population) | Italy | 1975-2003 | 0-28 years | 5209 men (8 suicide cases) | NR (9.92 expected) | Suicide: 0.81 (0.35, 1.59) |
| Fernandes et al. (2019),<sup>11</sup> retrospective cohort study | Former professional athletes (orthopaedic patients) | England | NR | NR | 572 men (33 depression 'caseness') | 500 (28 depression 'caseness') | Depression: 1.03 (0.61, 1.73) |
| Russell et al. (2020),<sup>8</sup> retrospective cohort study | Former professional athletes (general population) | Scotland | NR | Median 18 years | 7676 men (38 depression cases; 19 suicide cases) | 23028 males from the general population (169 depression cases; 93 suicide cases) | Depression: 0.64 (0.44, 0.92)<br>Suicide: 0.69 (0.25, 1.87) |
| **American Football** |  |  |  |  |  |  |  |
| Lehman et al. (2016),<sup>9</sup> retrospective cohort study | Former professional athletes (general population) | USA | 1959-1988 | Maximum 34 years | 3439 (12 suicide deaths) | NR (25.6 expected) | Suicide: 0.47 (0.24, 0.82) |
| Lincoln et al. (2018),<sup>29</sup> retrospective cohort study | Former professional athletes (general population) | USA | 1986-2012 | 2-28 years | 9778 (20 suicide deaths) | NR (34.5 expected) | Suicide: 0.58 (0.35, 0.90) |
| Phelps et al. (2022),<sup>15</sup> retrospective cohort study | Former high school athletes (general population) | USA | 1964-1980 | 38-55 | 216 (30 depression 'caseness') | 638 (94 depression 'caseness') | Depression: 0.93 (0.60, 1.45) |
| **Rugby union** |  |  |  |  |  |  |  |
| Decq et al (2016),<sup>30</sup> retrospective cohort study | Former amateur athletes (general population) | France | 1985-1990 | NR | 239 (21 depression 'caseness') | 138 (8 depression 'caseness') | Depression: 1.56 (0.67, 3.64) |

NR, not reported

**Figure 1.**
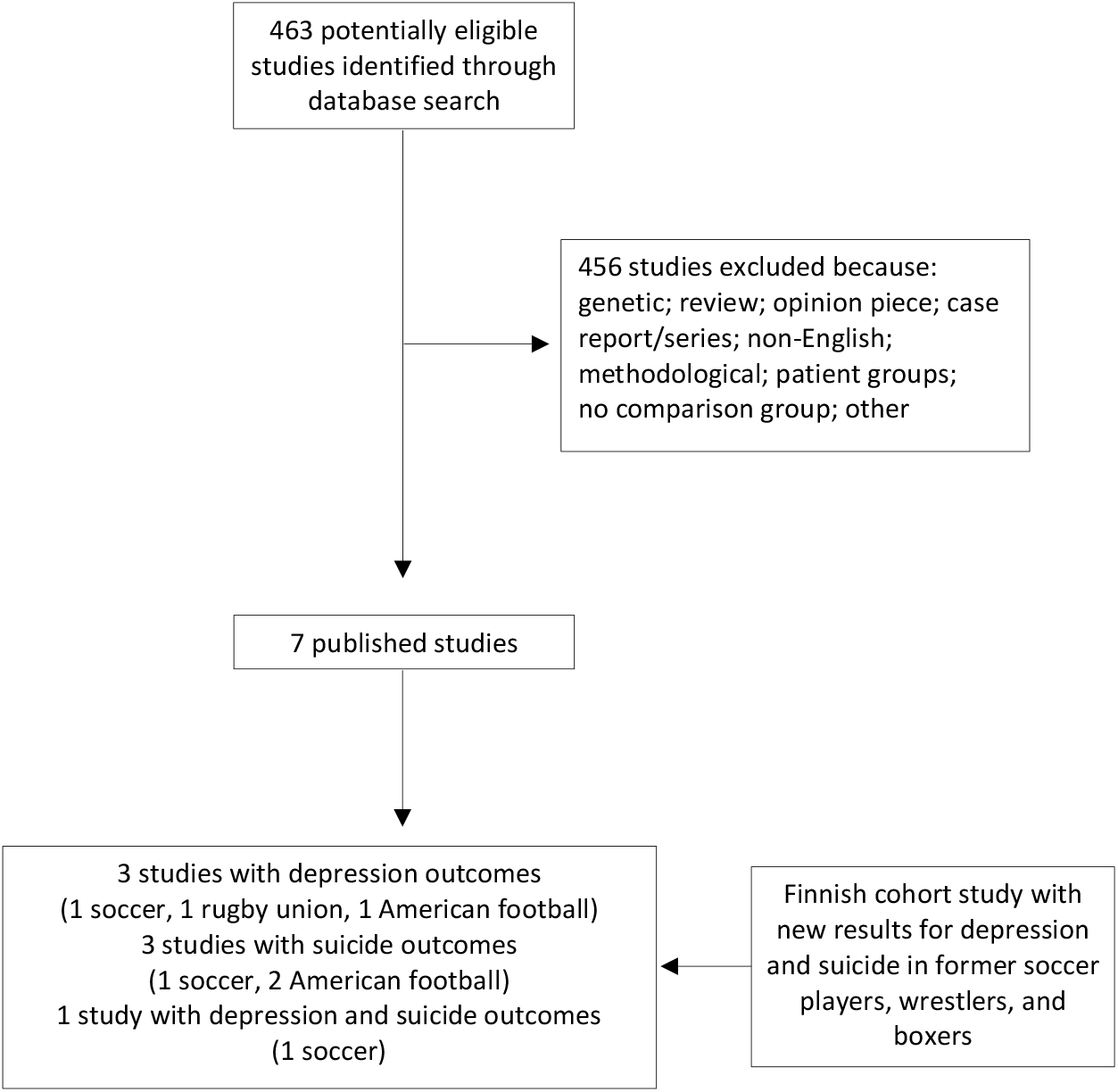
Study selection: Systematic review.

On combining effects estimates for all contact sports in the meta-analysis, including the results from the Finnish cohort (4 effect estimates), there was little evidence of an association with depression (summary risk ratio: 0.89 [95% CI 0.71; 1.10]). In corresponding analyses for suicide, contact sports participants experienced a lower risk (0.71 [0.52; 0.98]) relative to general population controls. Heterogeneity across these studies was low (depression: I^2^=10%, p-value=0.35; suicide: I^2^=19%, p-value=0.28). On stratifying according to study quality, higher- (≥7) and lower-scoring studies yielded similar results for both depression and suicide.

In analyses of specific sports, we aggregated results when there was a minimum of two studies capturing the same activity (figures 2 and 3). We found a lower risk of depression amongst former soccer players relative to control groups (4 studies: 0.71 [0.54, 0.93]; I^2^=0%, p-value=0.40). When we stratified according to studies sampling former professionals (2 studies: 0.78 [0.49, 1.24]; I^2^=53%, p-value=0.06)^8,11^ and amateurs in the Finnish cohort (2 studies: 0.57 [0.31, 1.04,]; I^2^=0%, p-value=0.69), there was a suggestion of a lower risk of depression in both groups but not at conventional levels of statistical significance. There was only one study of depression in retired American football players,^15^ and the prevalence of depression in this cohort of former high school participants was not appreciably different to that of the general population (table 3).

**Figure 2.**
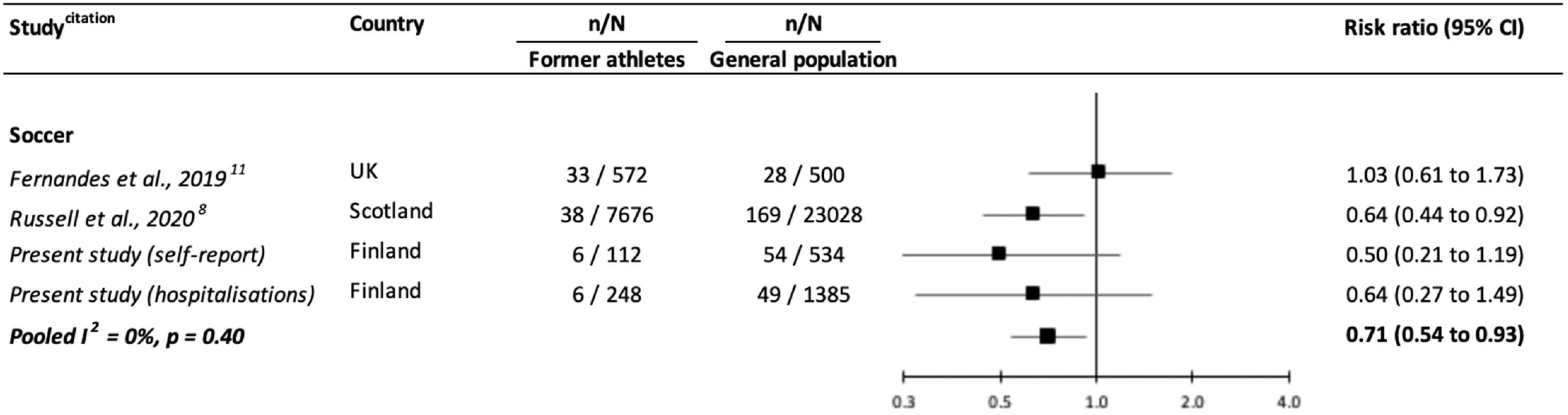
Risk ratios (95% confidence intervals) for the relation of former participation in soccer with depression: Meta-analysis.

**Figure 3.**
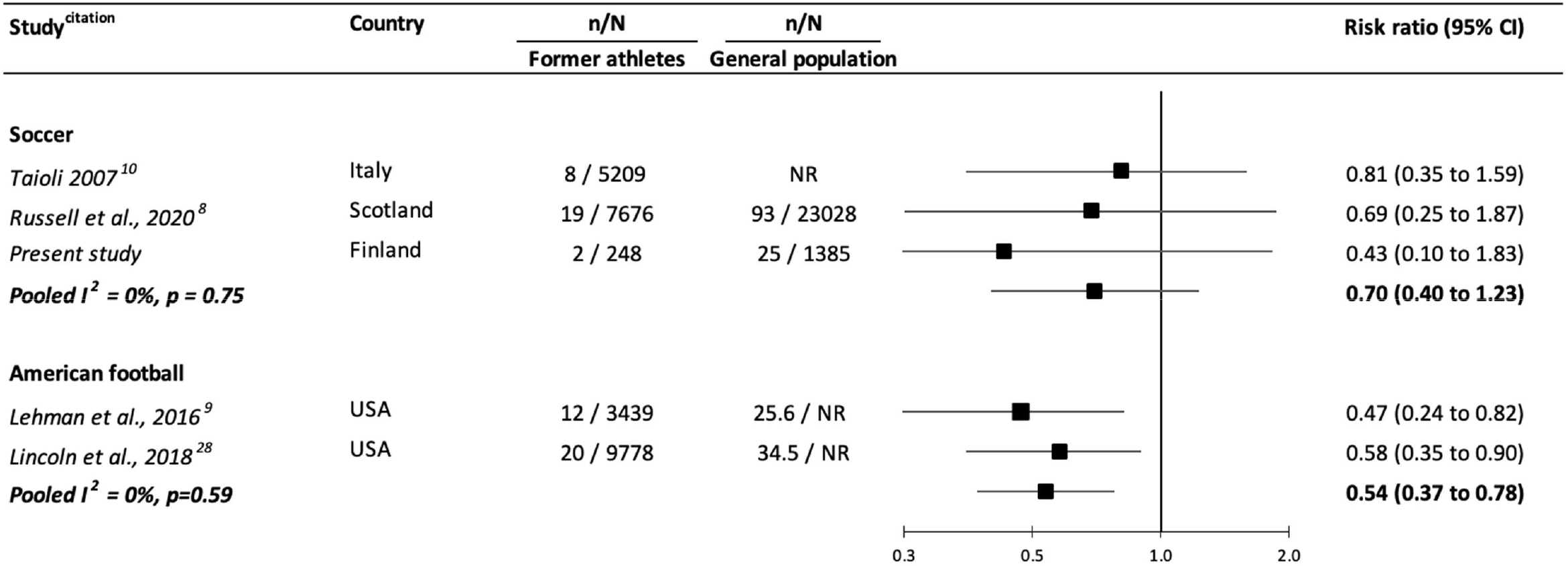
Risk ratios (95% confidence intervals) for the relation of former participation in contact sports with suicide: Meta-analysis.

In analyses of studies with data on suicide (figure 3), retired soccer players had somewhat lower rates than the general population but not significantly so (3 studies: 0.70 [0.40, 1.23]; I^2^=0%, p-value=0.75). There was no suggestion of a differential effect for former amateurs (0.43 [0.10, 1.83])^8,11^ versus professional players (2 studies: 0.76 [0.42, 1.40], I^2^=0%, p-value=0.80). In the two studies of retired American football players, however, a background in this collision sport was associated with protection against completed suicide (2 studies: 0.54 [0.37, 0.78]; I^2^=0%, p-value=0.59).

## Discussion

In the present report, we aggregated new results from analyses of a cohort study of former contact sports participants with those from the extant literature. With the caveats that the evidence base is modest in scale and confined to men, there was no suggestion that retired soccer and American football players had poorer mental health than the general population. Rather, we found that erstwhile soccer players appeared to have a lower risk of depression relative to general population controls, while former American football players appeared to experience some protection against suicide.

These epidemiological observations run counter to those made in case series of athletes from contact sports who, at autopsy, were found to be seemingly affected by a combination of chronic traumatic encephalopathy, depression, and suicidal tendencies,^5,6^ although this interpretation, particularly in the absence of an unexposed comparator group, has been challenged.^7^ In a separate body of literature, traumatic brain injury requiring hospitalisation has been linked to a greater future occurrence of depression and suicide in large scale cohort studies.^1,2^ It may be that head impact in the contact sports included herein are of insufficient severity to precipitate long-term depression and suicide. Alternatively, post-retirement level of physical activity in elite athletes is seemingly higher than the general population,^31^ and, as such, the apparent preventative effect of long-standing patterns of physical exertion against depression^32^ and suicidal ideation^33^ may be compensating for the deleterious effect, if any, of low level head impact.

### Study strengths and limitations

The present study has its strengths, including being the first synthesis of depression risk in former participants from contact sports, and one that incorporates new cohort study data. It is not, however, without its limitations. First, all included studies exclusively sample men. There is some evidence of sex differentials in other risk factors for depression^34^ and suicide,^35^ so the extent to which the present findings for soccer can be generalised to women is moot. Second, the findings of a meta-analysis are only as strong as the methodological quality of the studies it includes and, although half the studies were judged to be of high grade, all data were nonetheless observational. With conventional trials in this field being unviable ethically and perhaps logistically, an advance on current evidence may be the use of natural experiments. These could include the impact on depression or suicide risk pre- and post-introduction of compulsory protective equipment such as change in the composition of the soccer ball from leather to plastic which resulted in lower weight (1986-present), or the introduction of head gear in amateur boxing (1984-2016). Third, none of the included studies had data on actual head impacts; instead, sporting background was used as a proxy. There is empirical evidence, however, of a higher occurrence of head trauma in contact sports groups versus control populations.^36^

In conclusion, based on a modest number of studies exclusively comprising men who were largely from professional backgrounds, retired soccer players had a lower risk of depression and former American football players had a lower risk of suicide at follow-up. Whether these findings are generalisable to women requires testing.

## Data Availability

Interested parties should contact UMK and SJS regarding access to the Finnish cohort study data.

## Acknowledgement

The preparation of this manuscript received no direct funding. GDB is supported by the UK Medical Research Council (MR/P023444/1) and the US National Institute on Aging (1R56AG052519-01; 1R01AG052519-01A1); PF by the UK Economic and Social Research Council & Biotechnology and Biological Sciences Research Council (Soc-B Centre for Doctoral Training); and JK by the Academy of Finland Centre of Excellence in Complex Disease Genetics (336823).

## Supplemental file

### Supplemental box 1. Search string for Pubmed

The PROSPERO (CRD42022352780) entry describes a series of health outcomes. The search here was specifically for depression and suicide.

(boxing[MeSH] OR boxing[title/abstract] OR boxer[title/abstract] OR boxer s[title/abstract] OR boxers[title/abstract] OR martial[title/abstract] OR wrestl*[title/abstract] OR football[MeSH Terms] OR football[title/abstract] OR footballers[title/abstract] OR footballs[title/abstract] OR soccer[MeSH Terms] OR soccer[title/abstract] OR hockey[MeSH Terms] OR hockey[title/abstract] OR rugby[MeSH Terms] OR rugby[title/abstract] OR racquet sports[MeSH Terms] OR racquet[title/abstract] OR racquet sports[title/abstract] OR lacrosse[title/abstract])

AND

(depression[title/abstract] OR depressive[title/abstract] OR depression[Mesh] OR depressive disorder[Mesh:NoExp] OR major depressive disorder[Mesh] OR dysthymic disorder[Mesh] OR Suicid*[Title/Abstract] OR suicide[Mesh])

**Supplemental Table 1.**
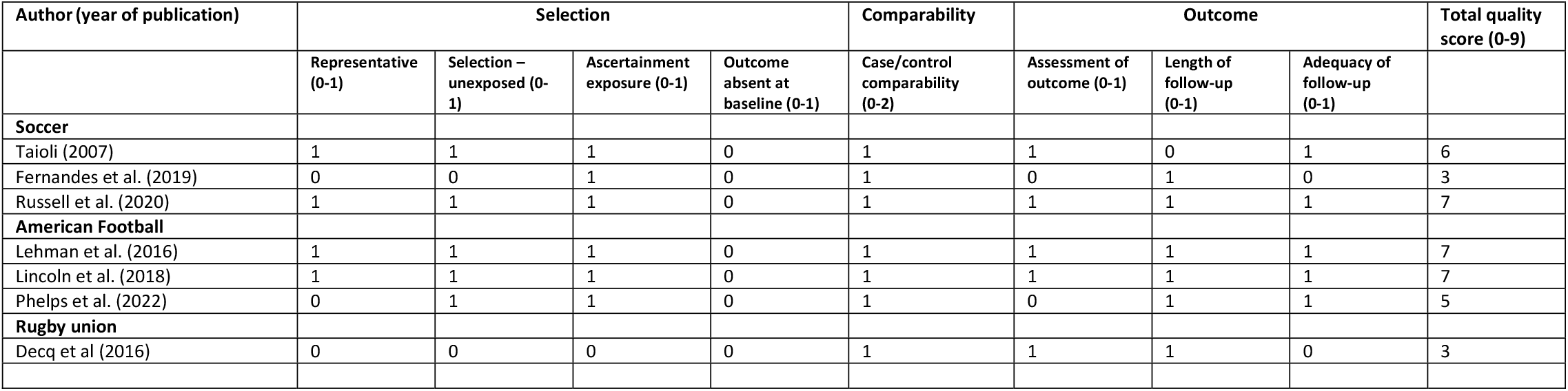
Cohort study quality assessment according to the Newcastle-Ottawa criteria: Meta-analysis.

**Supplemental table 2.**
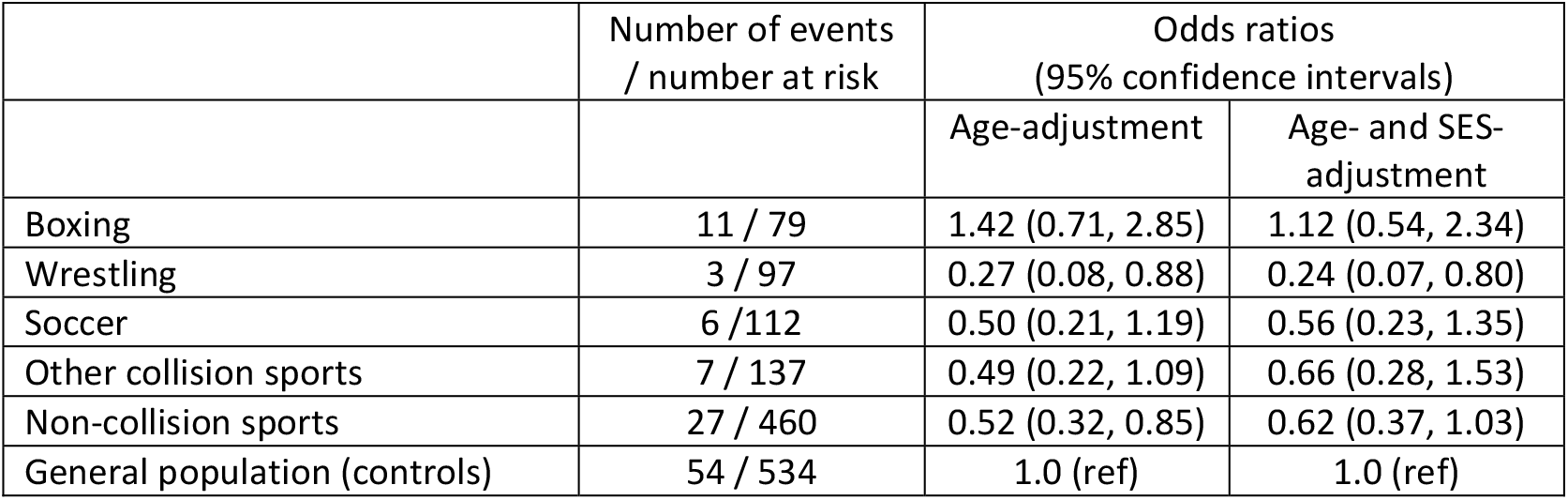
Odds ratios (95% confidence intervals) for the association of participation in contact sports with depression ‘caseness’: Finnish cohort study.

## Notes

Conflict of interest: None to declare.

### Competing Interest Statement

The authors have declared no competing interest.

### Author Declarations

Data collection was approved by the ethics committee of the Hospital Districts of Helsinki and Uusimaa, and all participants consented.

