## supplemental file for "Suicide and depression in former contact sports participants: population-based cohort study, systematic review, and meta-analysis"

**Supplemental box 1. Search string for Pubmed**

The PROSPERO (CRD42022352780) entry describes a series of health outcomes. The search here was specifically for depression and suicide.

(boxing[MeSH] OR boxing[title/abstract] OR boxer[ title/abstract] OR boxer s[title/abstract] OR boxers[title/abstract] OR martial[title/abstract] OR wrestl*[title/abstract] OR football[MeSH Terms] OR football[title/abstract] OR footballers[title/abstract] OR footballs[title/abstract] OR soccer[MeSH Terms] OR soccer[title/abstract] OR hockey[MeSH Terms] OR hockey[title/abstract] OR rugby[MeSH Terms] OR rugby[title/abstract] OR racquet sports[MeSH Terms] OR racquet[title/abstract] OR racquet sports[title/abstract] OR lacrosse[title/abstract])

AND

(depression[title/abstract] OR depressive[title/abstract] OR depression[Mesh] OR depressive disorder[Mesh:NoExp] OR major depressive disorder[Mesh] OR dysthymic disorder[Mesh] OR Suicid*[Title/Abstract] OR suicide[Mesh])

**Supplemental Table 1. Cohort study quality assessment according to the**

**Newcastle-Ottawa criteria: Meta-analysis**

| **Author (year of publication)** | **Selection** | | | | **Comparability** | **Outcome** | | | **Total quality score (0-9)** |
| --- | --- | --- | --- | --- | --- | --- | --- | --- | --- |
|  | **Representative(0-1)** | **Selection – unexposed (0-1)** | **Ascertainment exposure (0-1)** | **Outcome absent at baseline (0-1)** | **Case/control comparability**  **(0-2)** | **Assessment of outcome (0-1)** | **Length of follow-up**  **(0-1)** | **Adequacy of follow-up**  **(0-1)** |  |
| **Soccer** |  |  |  |  |  |  |  |  |  |
| Taioli (2007) | 1 | 1 | 1 | 0 | 1 | 1 | 0 | 1 | 6 |
| Fernandes et al. (2019) | 0 | 0 | 1 | 0 | 1 | 0 | 1 | 0 | 3 |
| Russell et al. (2020) | 1 | 1 | 1 | 0 | 1 | 1 | 1 | 1 | 7 |
| **American Football** |  |  |  |  |  |  |  |  |  |
| Lehman et al. (2016) | 1 | 1 | 1 | 0 | 1 | 1 | 1 | 1 | 7 |
| Lincoln et al. (2018) | 1 | 1 | 1 | 0 | 1 | 1 | 1 | 1 | 7 |
| Phelps et al. (2022) | 0 | 1 | 1 | 0 | 1 | 0 | 1 | 1 | 5 |
| **Rugby union** |  |  |  |  |  |  |  |  |  |
| Decq et al (2016) | 0 | 0 | 0 | 0 | 1 | 1 | 1 | 0 | 3 |

|  | Number of events / number at risk | Odds ratios  (95% confidence intervals) | |
| --- | --- | --- | --- |
|  |  | Age-adjustment | Age- and SES-adjustment |
| Boxing | 11 / 79 | 1.42 (0.71, 2.85) | 1.12 (0.54, 2.34) |
| Wrestling | 3 / 97 | 0.27 (0.08, 0.88) | 0.24 (0.07, 0.80) |
| Soccer | 6 /112 | 0.50 (0.21, 1.19) | 0.56 (0.23, 1.35) |
| Other collision sports | 7 / 137 | 0.49 (0.22, 1.09) | 0.66 (0.28, 1.53) |
| Non-collision sports | 27 / 460 | 0.52 (0.32, 0.85) | 0.62 (0.37, 1.03) |
| General population (controls) | 54 / 534 | 1.0 (ref) | 1.0 (ref) |

**Supplemental table 2. Odds ratios (95% confidence intervals) for the association of**

**participation in contact sports with depression ‘caseness’: Finnish cohort study**
